# Effectiveness of a SARS-CoV-2 mRNA vaccine booster dose for prevention of infection, hospitalization or death in two nation-wide nursing home systems

**DOI:** 10.1101/2022.01.25.22269843

**Authors:** Kevin W. McConeghy, Barbara Bardenheier, Andrew W. Huang, Elizabeth M. White, Richard A. Feifer, Carolyn Blackman, Christopher M. Santostefano, Yoojin Lee, Frank DeVone, Christopher W. Halladay, James L. Rudolph, Andrew R. Zullo, Vincent Mor, Stefan Gravenstein

## Abstract

**Background:** A SARS-CoV-2 vaccine booster dose has been recommended for all nursing home residents. However, we lack effectiveness data on boosters preventing infection, death and hospitalization in this frail population.

**Methods:** We emulated nested target trials in two large nursing home systems in parallel to evaluate the effectiveness of a SARS-CoV-2 mRNA vaccine booster at preventing infection, hospitalization, or death. Residents who completed a 2-dose series of the mRNA vaccine and were eligible for a booster were included in from September 22, 2021 to November 5, 2021. Outcomes were measured through December 18, 2021, including test-confirmed SARS-CoV-2 infection, hospitalization, or death. The vaccine effectiveness at day 42 was estimated with a Kaplan-Meier estimator, both unadjusted and weighted with the inverse probability of treatment.

**Results:** The two NH systems were large and multi-state, System 1 included 200 NH (8,538 control and 5,721 boosted residents) and System 2 included 127 NHs (4,100 control and 2,291 boosted residents). Booster vaccination reduced infections by 50.4% (95% Confidence Interval [CI]: 29.4%, 64.7%) SARS-CoV-2 infections in System 1 and 58.2% (32.3%, 77.8%) in System 2. Boosted residents in System 1 also had a 97.3% (86.9%, 100.0%) reduction in SARS-CoV-2 associated death, but too few events for comparison in System 2.

**Conclusions:** During a Delta predominant period, SARS-CoV-2 booster vaccination significantly reduced infection in two U.S. nursing home systems. In the larger System 1 a 97% reduction in SARS-CoV-2 related death was also observed. These findings strongly support administration of vaccine boosters to nursing home residents.

## Introduction

Nursing home residents have suffered significant SARS-CoV-2 morbidity and mortality.^1,2^ Resurgence in late summer/early fall 2021 with the Delta variant and waning immunity in NURSING HOME residents^3^ led the Centers for Disease Control and Prevention (CDC) to recommend an additional “booster” dose for those vaccinated with a primary series.^4,5^ The Food and Drug Administration amended the emergency use authorizations for the BNT162b2 (Pfizer-BioNTech) and mRNA-1273 (Moderna) vaccines in September and October 2021, respectively, to include an additional dose at least six months after the primary series for ‘high-risk’ individuals, including those aged 65 years or older.^6^ Both mRNA vaccines were also authorized in August 2021 to be used for additional doses in immunocompromised individuals, a description that applies to many long-term care residents. As of December 12, 2021, approximately 55% of U.S. nursing home residents had received a booster.^7^

Despite the high risk of SARS-CoV-2 morbidity and mortality in nursing home residents, limited data exist on the effectiveness of boosters in mitigating infection or death in this frail population. Vaccine trials did not include nursing home residents, and timely patient-level observational data are not widely available. Nursing homes are an optimal environment for measurement of vaccine effectiveness because of the residential stability, systematic documentation of immunizations (including boosters) and frequent testing for SARS-CoV-2. Additional clinical evidence of booster effectiveness would support efforts to increase booster distribution for this vulnerable population.

To address this gap, we leveraged electronic health record data updated daily from two large, multi-state long-term care providers. Using a nested target trial emulation method^8^, we evaluated the vaccine effectiveness of an mRNA booster dose (versus no booster) among residents who completed an mRNA vaccination primary series. Our outcomes included any laboratory-confirmed SARS-CoV-2 infection, hospitalization, and death. We hypothesized that a mRNA vaccine booster would prevent these outcomes.

## Methods

### Study Design and Population

This study was designed to emulate a sequence of nested target trials comparing additional effectiveness of SARS-CoV-2 mRNA vaccine booster versus primary series only in nursing home residents. The first system included 200 nursing homes in 19 states with a heavy concentration in the northeast region operated by Genesis HealthCare (System 1). The second included Veterans residing in 127 community living centers (analogous to nursing homes) spread nationwide managed by the Veterans Health Administration (System 2). The study emulates daily (Monday – Friday) target trials beginning September, 22, 2021 through November 5, 2021. September 22 was the date that the FDA authorized the BNT162b2 vaccine booster in nursing home residents. November 5, 2021 was selected as the last trial day allowing at least 6 weeks follow-up observation time to December 18^th^, 2021. Data was collected from administrative and clinical records in either system. This study was deemed to be exempt by the Brown University institutional review board (IRB) and the VA data was approved by the Providence VA Medical Center IRB.

The emulated target trial method involves designing a hypothetical trial and emulating it in the observational data through applying inclusion and exclusion criteria to a cohort, and identifying a well-defined intervention and outcome.^8,9^ Specific dates are selected as ‘trial’ dates where those meeting eligibility and assigned to treatment or not are eligible for inclusion. Our target trial is outlined in Table 1 for both nursing home systems. This represents the trial being emulated in the observational data. On each day residents were considered eligible for inclusion if they had been present in the home for at least 90 days with receipt of a 2-dose mRNA vaccine series at least 180 days earlier. Residents who received the Janssen vaccine as their initial vaccination were excluded due to small numbers. Residents were also excluded if they had a SARS-CoV-2 infection in the prior 90 days, planned discharge according to their most recent assessment, received monoclonal antibodies in the prior 90 days, were under hospice care, or had already received a third vaccine dose prior to the index date.

| Component | Target Trial | Cohort 1: Community nursing home | Cohort 2: VA nursing home |
| --- | --- | --- | --- |
| **Aim** | Relative risk for SARS-CoV-2 infection with booster vaccination versus primary series only |  |  |
| **Eligibility** | **Inclusion**<br>Present in nursing home on any index date (09/22 – 11/5/21)<br>Admitted at least 90+ days prior to index<br>Received full mRNA primary vaccination series (two doses + 14 days) at least 180 days prior to index<br>**Exclusion**<br>A medical history of SARS-CoV-2 within 30 days prior to index (any ICD code / (+) PCR)<br>Treatment with monoclonal antibody within 90 days prior to index<br>Already received SARS-CoV-2 booster prior to index date<br>Hospice care | **Additional:**<br>☐ complete case analysis (excluding missing data)<br>☐ Nursing home operated by Genesis as of 12/18/2021<br>☐ No plan for discharge | **Additional:**<br>Allow 1-day gap in in 90 days prior |
| **Treatment** | Receipt of COVID-19 booster dose (third dose more than six months after primary series) compared to placebo | Instead of placebo, we compare to individuals without a booster | Same |
| **Assignment** | Random assignment by individual | Non-randomized, use inverse-probability treatment weighting to adjust for selection bias | Same |
| **Follow-up** | **Censoring events**<br>Death<br>Transfer out of nursing home (hospital or other) | Also: receipt of booster (in control arm) | Same; transfer out of VA institutional care (i.e. to community) |
| **Outcome** | SARS-CoV-2 (+) PCR/Antigen result along with SARS-CoV-2 associated events: infection, hospitalization or death | Same | Same |
| **Statistical Analysis** | Kaplan-Meier estimator, cumulative incidence, vaccine effectiveness at specific timepoints, day 42 (6 weeks) | Same | Same |
| **Description.** This table outlines the hypothetical design of a randomized trial in nursing home residents, defining the intervention, inclusion/exclusion criteria and outcomes. The right-most columns highlights where the emulated data for each cohort deviates from this design. |  |  |  |

### Study Outcomes and Covariates

We examined four post-booster outcomes: (1) any incident SARS-CoV-2 infection; (2) hospitalization (3) death, and (4) a combined endpoint of hospitalization or death. All study outcomes were obtained via the electronic health record from the two systems.

**SARS-CoV-2 infection** was defined by a positive SARS-CoV-2 rapid antigen or polymerase chain reaction test occurring after the index trial date, and with no test confirming infection in the prior 90 days. Testing results were extracted from the laboratory results of either system.^10^

**SARS-CoV-2 associated hospitalization** defined as transfer to an acute care hospital occurring within 21 days of (+) SARS-CoV-2 test. Transfers were identified through census records in system 1 and bed-section codes identifying acute care episodes in system 2, which has a single EHR system for both nursing homes and hospitals.

**SARS-CoV-2 associated death** defined as death occurring within 30 days of a (+) SARS-CoV-2 test. Deaths in System 1 were identified through transfers to funeral homes, or death records in the daily census. System 2 deaths were identified through a combination of census and vital status records.

Immunization records in each EHR were used to identify primary vaccination and booster events.^11^ We indexed additional resident covariates separately for each eligible trial date (i.e. a person’s baseline covariates may differ for each trial, as each trial represents a different index date). Data was obtained from the electronic health record in both systems. In system 1, additional information from the nursing home minimum dataset assessments (MDS) were included.^12^ MDS includes demographic characteristics (age, sex, race/ethnicity, language), clinical diagnoses, MDS measures of cognitive and physical function, an MDS indicator of limited life expectancy, prior SARS-CoV-2 infection, and influenza vaccination for the current season up to the index date. System 2 data did not include minimum dataset assessments and relied on demographics and clinical diagnoses codes in the EHR (recorded in the past 12 months prior to index) for chronic condition information. These were classified using the Elixhauser comorbidity classification system.^13^ We characterized residents as immunocompromised if they had received an immunosuppressive medication or a qualifying condition diagnosis (detailed in supplementary appendix Table S1). Finally, we included measures of overall facility testing SARS-CoV-2 positivity rates in the 14-day period prior to each index date.

### Statistical analysis

#### Nested trial method

The analytic approach follows a general daily nested target trial methodology.^8^ The purpose of the approach was to provide a flexible method for assigning “time-zero” to unboosted residents, while being maximally inclusive of follow-up data for unboosted residents (up to the time they are boosted). When a resident receives a booster vaccination on a trial date, they are assigned to the booster arm.

Once a resident is boosted, they are ineligible for future trial dates, and therefore the booster arm does not contain repeated observations. Unboosted residents might be eligible as controls for multiple emulated trial dates up until they received a booster. We randomly selected one eligible target trial date per control resident.^8^ From each trial date, a ‘survival’ time was computed as the minimum of either time to outcome, or a censoring event (i.e. transfer from facility, death, or last available date of follow-up [December, 18, 2021]). Controls were censored if they subsequently receive a booster dose, but contributed follow-up time until the point of censoring on the date of booster administration.

### Statistical Analysis

Both cohorts used the same analytical approach. Each event was modeled using the Kaplan-Meier estimator, with cumulative incidence (i.e., risk) measured as 1 - probability of event free survival at each time point (day). The relative ratio of the cumulative incidence curves between groups (boosted, unboosted) at each time point was computed, with 1 - relative risk reported as the conventional “vaccine effectiveness” statistic. The cumulative incidence at day 42 (six weeks) was selected as the primary follow-up time of interest for reporting. We conducted a complete case analysis and did not impute missing data as the number of observations excluded for missing data was very small (<1% observations dropped). Final statistical models were selected by comparison of covariate imbalances between groups or *a priori* based on experience (i.e. gender, facility-level infection rates). We estimated a stabilized inverse probability weight for treatment (IPTW) adjusting for: the facilities’ prior 90-day hospitalization rate, restricted cubic splines for the number of resident-level SARS-CoV-2 tests, and facility positivity rate in the prior 14 days, state (System 1 only), primary vaccination series manufacturer, month of trial date (September versus October, November), resident immunocompromised status, age, and race/ethnicity. System 2 also adjusted for a diagnosis of paralysis, alcohol, weight-loss and blood-loss. These variables were selected by comparing between groups using standardized mean differences (SMD), with an absolute SMD<0.1 after adjustment as a guide for achieving acceptable balance between groups. Weights were truncated at their 99% upper quantile. Sampling with replacement by resident (i.e. bootstrapping) with 2000 replications was used to generate 95% percentile intervals accounting for the estimated probability weights and crossover of treatment groups by resident. Initial data preparation was done with SAS Version 9.4 (Cary, NC) and STATA version 16 (Statacorp, TX). All analyses were performed using R statistical software, Version 4.0.1, Vienna, Austria.

## Results

### Study Population

Tables S2, S3 and Figure S3 in the supplementary appendix provide resident counts by inclusion and exclusion criteria for each day.

System 1 (Genesis) included 8,587 unique residents across 200 nursing homes. 8,538 served as controls for at least 1 day of follow-up and 5,672 residents received a booster during the study period and were eligible to be included in the boosted arm. There were 5,624 residents present in both arms of the study first as controls, then the boosted arm.

System 2 (VA) included unique 6,391 residents across 127 nursing homes. 4,100 served as controls for at least 1 day of follow-up and 2,291 residents received a booster at some point during the study period and were eligible to be included in the boosted arm. There were 2,283 residents present in both arms of the study first as controls, then boosted residents.

In both systems, the majority of residents received a Pfizer-BioNTech booster 4,259 (75.1%) and 1,276 (55.7%), respectively. The follow-up time contributed was 557,817 days (median 31 days per resident, Q1:11, Q3:66) in System 1 and 229,281 (median 35 days per resident, Q1:10, Q3:53) in System 2.

Table 1 provides baseline characteristics for the larger cohort from System 1 including the boosted and unboosted (control) groups. A similar table for System 2 is available in Supplementary Table S4. There were some significant differences between systems, System 1 was older (78 versus 72 years), had fewer black residents (10% versus 25%) and more female residents (64% versus 4%). The two treatment groups were similar (standardized mean differences <0.1) by most observed characteristics in both System cohorts. The primary difference between treatment groups observed was at the facility level, where unboosted controls were more likely than boosted to be facilities with a SARS-CoV-2 infection in the prior 14 days (17.3% vs. 11.2%, aSMD 0.17). Supplementary figures S4 and S5 describe the propensity score and treatment weights.

### Effects of Booster Dose

In Figure 2 we present cumulative incidence curves for the four COVID-19 outcomes up to 6 weeks (day 42) for System 1. Table 3 summarizes vaccine effectiveness observed in both systems. Booster vaccination reduced risk for infection 50.4% (95% Confidence Interval [CI]: 29.4%, 64.7%) in System 1 and 58% (32.3%, 77.8%) in System 2, respectively. SARS-CoV-2 hospitalization was lower in boosted groups (VE 47.7% and 36.6% respectively), but a null effect could not be excluded in IPTW-adjusted analyses. In System 1, there was a 97.3% (86.9%, 100.0%) VE for SARS-CoV-2 associated death, 1 in boosted versus 15 deaths in unboosted. In System 2, there were 2 deaths in the boosted arm and 3 in the unboosted arm. The limited number of events made a confidence interval difficult to estimate so VE was not reported. VE for a combined endpoint of death or hospitalization was 82.0% (55.5%, 94.0%) in System 1 versus 45.8% (−15.5%, 79.1%) in System 2.

## Discussion

This study evaluated the clinical effectiveness of an mRNA SARS-CoV-2 vaccine booster in two large multi-state populations of nursing home residents. We sought to estimate average booster vaccine effectiveness comparable to trial eligibility criteria. We found that those who received a booster dose had a substantial reduction in risk of SARS-CoV-2 infection and death in System 1, while in System 2 there was a similar reduction in infection but we were unable to exclude null effects for other endpoints. In the larger System 1, one SARS-CoV-2 associated death occurred following boosting, versus 15 deaths in non-boosted individuals, but smaller sample size and event counts made inference on death events in System 2 difficult. While some event rates are low, and magnitude of effects differ, the findings between cohorts are overall consistent in that those receiving boosters had a significant reduction in risk of SARS-CoV-2 infection.

To evaluate the robustness of vaccine effectiveness, we examined two significantly different nursing home populations; one a large, private U.S. provider of nursing home care and the other a public, national healthcare system caring specifically for elderly Veterans. These two health systems are demographically different (e.g. few women are present in VA CLCs), and represent two different administrative healthcare systems. In particular, the rates and reasons for acute hospitalization may be different between two populations due to differences in availability of hospice care, advanced planning and provider practice. It should also be noted the sample size for the VA CLCs (System 2) is considerably smaller (approximately half the residents of System 1), with less follow-up time and fewer facilities. These differences mean direct comparisons of the VE or pooling of results is inappropriate. However, consistency in overall vaccine effectiveness to prevent infections in these two disparate cohorts lends credibility to the importance of boosters for the overall U.S. nursing home population.

There are few other studies of booster effectiveness in nursing home residents to date. Bar-On et al reported observational data in Israeli community-dwelling adults aged 60 and older boosted 5 months after primary vaccinations.^14^ Comparing confirmed infections and adjusting for person-days at risk, older adults had about 88% fewer confirmed infections in the 12 to 42 days following the boost when the Delta variant was the predominant virus in circulation. In a second similar approach for comparison, Israelis aged 16 and older followed from 12 to up to 60 days following boost, experienced 9 to 17-fold fewer infections in person days at risk.^15^ From the same period, Muhsen at al reported on over 41,000 Israeli nursing home residents aged 60 years and older boosted ∼5 months after the primary series in a three-week campaign.^16^ In that analysis, the contemporary comparison included 1.5 million community dwelling adults 60 and older, and younger groups aged 20-59 years and under 20 years. The nursing home residents had an average age of 81.8, but the average age of the 60 and older comparison group was not provided. Nonetheless, the boosted nursing home residents had a 71% lower rate of infections 80% lower hospitalizations than their unboosted community dwelling contemporaries. These ecological analyses suggest substantial protection by boosting against infection, hospitalization and death, but cannot account for confounding by access, indication or exposure risk in the outpatient comparison populations. Indeed, the outpatient elderly group reportedly had less risk reduction than the nursing home resident group, as did the younger comparison groups.

The present work evaluates VE by emulated trial design that takes advantage of comparable controls who differ primarily by when they received vaccine rather than by indication, exposure risk to circulating strains, and residents in facilities across a broad geographic and socioeconomic distribution. We report a slightly lower VE of 50-54% infection reduction comparing boosted and unboosted residents, and included all post-booster days in the exposure risk calculation during which boost-derived immunity may be incomplete. Also, the VE to prevent SARS-CoV-2 mortality in System 1 nursing home residents is substantially larger than reported by the Israeli studies, exceeding 90%. These differences could arise from differences in methods, population health, virus characteristics or other causes, but most likely from the first two.

Our study includes several significant limitations. To identify treatment effects, we assumed no unmeasured confounding and correct specification of probability weights in the Kaplan-Meier estimator. The relatively small differences between groups are encouraging and may arise from subjects’ eligibility requirement for prior primary series vaccination, resulting in greater cohort homogeneity. Additionally, the plots of our cumulative incidence curves for infection show close overlap in the first 7-10 days for groups. This suggests that we face relatively low risk of residual confounding if the two groups have similar baseline event risk in the window *after vaccination but prior to the quickly developing post-booster immunity*. A significant limitation that we do not account for is interference, or the assumption that one resident’s vaccination does not impact the effectiveness of vaccination for other residents. Subsequent work which accounts for this could lead to significantly different estimates as increasing proportions of boosted residents and staff protect unboosted residents from infection within a facility. Treatment effects assuming no interference likely underestimate the total causal effect. Additionally, deaths in the nursing home are easily identified, but for residents who are transferred, they may die after they are lost to follow-up, in which case we cannot account for them in our data.

This study reports a substantial reduction in risk of SARS-CoV-2 infection in the early phase of booster adoption by two large U.S. nursing home systems. These results strongly support vaccination with a SARS-CoV-2 booster dose for the entire eligible nursing home population.

## Data Availability

The data presented in this study contains protected health information and cannot be shared in its person-level form. However the authors can commit to responding to reasonable inquires about the data and sharing summary-level information when requested.

## Acknowledgements

The authors would like to acknowledge and thank Roee Gutman and Issa Dahebreh for providing advice on the statistical analysis plan. The authors would also like to thank John Jernigan, Stephanie Schrag and Jennifer Verani at the Centers for Disease Control and Prevention for provide guidance on study design and input during the analytical process. Additionally, we would like to thank Denine Hastings, Cliff Boyd and Jeff Hiris for providing support and assistance with data infrastructure and preparation of data files.

## Author Contributions

KWM conceived the main analytical approach and performed the primary analysis for the private nursing home system. BB, ARZ, EMW, VM, SG contributed to study design, interpretation of results and assisted in editorial review of the paper. AWH, CMS, YL assisted in preparing private nursing home data for analysis. RAF, CB provided and helped interpret the private nursing home data while also providing editorial review of the manuscript. JLR, FD, CAH provided the Veterans Health Administration analysis with oversight by KWM.

## Figures

**Figure 1.**
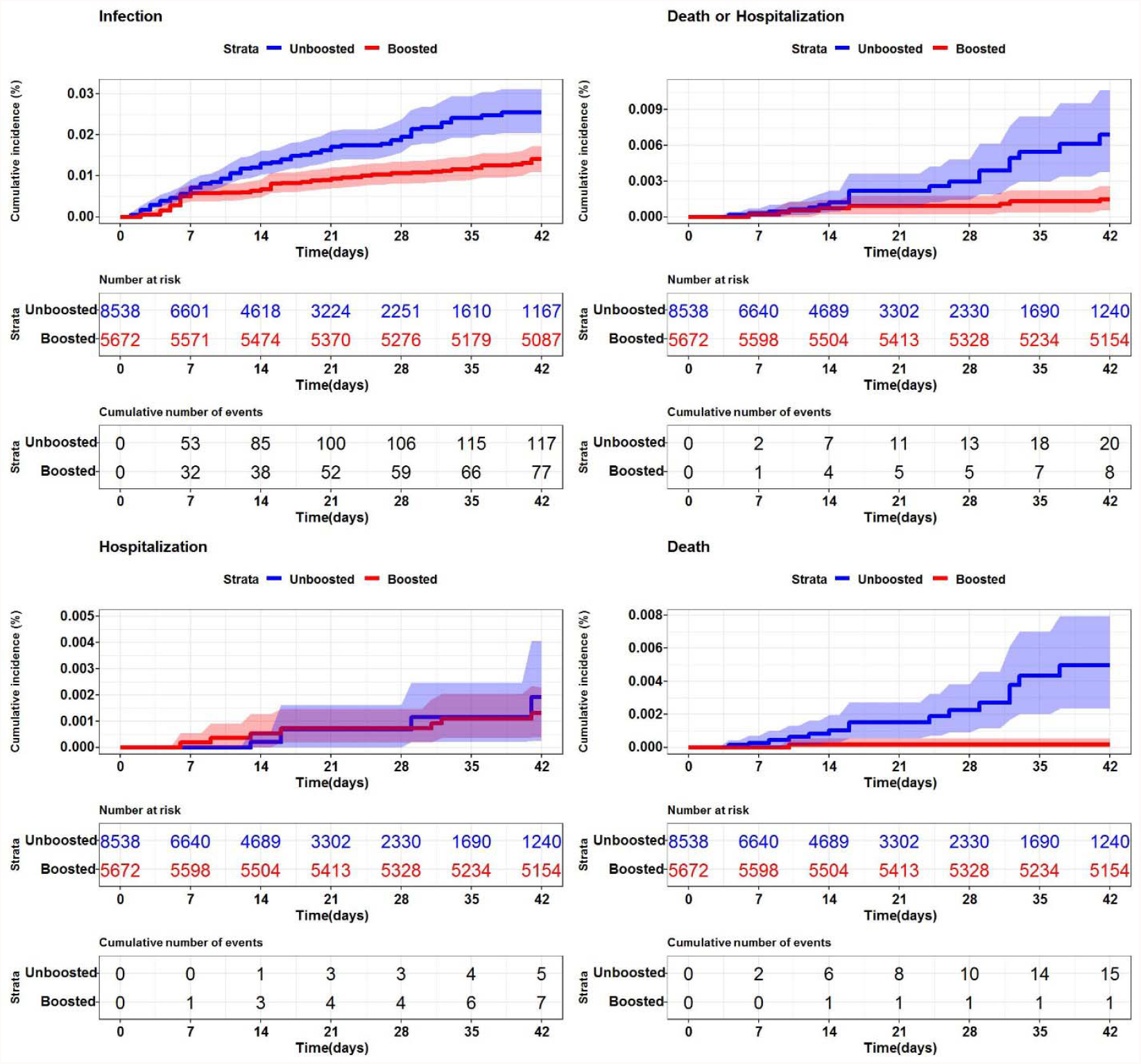
SARS-CoV-2 cumulative incidence in U.S. nursing homes (n=200) by booster status **Description.** Each graph represents the cumulative incidence for a different SARS-CoV-2 (+) outcome. (Top left - SARS-CoV-2 infection (any positive), Top right - SARS-CoV-2 death or hospitalization, Bottom left - SARS-CoV-2 associated hospitalization, Bottom right - death). The shaded regions represent 95% confidence intervals.

## Tables

**Table 2.** Baseline characteristics by booster status for private nursing home chain (System 1)

| Variable | Total<br>(n=14,210) | Unboosted<br>(n=8,538) | Boosted<br>(n=5,672) | aSMD |
| --- | --- | --- | --- | --- |
| Age, median (Q1, Q3) | 78 (69, 87) | 78 (68, 86) | 79 (69, 87) | 0.06 |
| Male, no. (%) | 5046 (35.5%) | 3093 (36.2%) | 1953 (34.4%) | 0.04 |
| Length of stay in days, median (Q1, Q3) | 796 (473, 1341) | 784 (460, 1326) | 819 (493, 1372) | 0.03 |
| Black, no. (%) | 1449 (10.2%) | 887 (10.4%) | 562 (9.9%) | 0.02 |
| Hispanic, no. (%) | 710 (5.0%) | 452 (5.3%) | 258 (4.5%) | 0.03 |
| Serious mental illness or mental disability, no. (%)<sup>a</sup> | 1232 (8.7%) | 778 (9.1%) | 454 (8.0%) | 0.04 |
| Current smoker, no. (%) | 467 (3.3%) | 306 (3.6%) | 161 (2.8%) | 0.04 |
| Needs language translator, no. (%) | 332 (2.3%) | 208 (2.4%) | 124 (2.2%) | 0.02 |
| Influenza vaccinated, no. (%)<sup>b</sup> | 6855 (48.2%) | 4040 (47.3%) | 2815 (49.6%) | 0.05 |
| SARS-CoV-2 history, no. (%) | 6848 (48.2%) | 4125 (48.3%) | 2723 (48.0%) | 0.01 |
| Dialysis/ESRD, no. (%) | 216 (1.5%) | 137 (1.6%) | 79 (1.4%) | 0.02 |
| Immunocompromised, no. (%) | 1637 (11.5%) | 961 (11.3%) | 676 (11.9%) | 0.02 |
| Pulmonary disease, no. (%) | 3131 (22.0%) | 1897 (22.2%) | 1234 (21.8%) | 0.01 |
| Diabetes mellitus (any), no. (%) | 5471 (38.5%) | 3284 (38.5%) | 2187 (38.6%) | <0.01 |
| Cognitive-function scale score, median (Q1, Q3)<sup>c</sup> | 2 (1, 3) | 2 (1, 3) | 2 (1, 3) | 0.01 |
| AHRQ chronic conditions, median (Q1, Q3) | 4 (3, 6) | 4 (3, 6) | 4 (3, 6) | 0.01 |
| DNR directive, no. (%)<sup>d</sup> | 7283 (51.3%) | 4306 (50.4%) | 2977 (52.5%) | 0.04 |
| Life expectancy less than six months, no. (%)<sup>e</sup> | 72 (0.5%) | 44 (0.5%) | 28 (0.5%) | <0.01 |
| Resident SARS-CoV-2 tests (past 14 days), mean (SD)<sup>f</sup> | 0.6 (1.8) | 0.6 (1.8) | 0.6 (1.7) | 0.03 |
| Resident SARS-CoV-2 tests (past 90 days), mean (SD)<sup>f</sup> | 3.3 (8.4) | 3.2 (8.0) | 3.6 (9.0) | 0.04 |
| Facility, any SARS-CoV-2 (+) in prior 14 days | 2116 (14.9%) | 1479 (17.3%) | 637 (11.2%) | 0.17 |
| **Description.** aSMD – absolute standardized mean difference, Q1/Q3 – 25/75<sup>th</sup> quartiles, SD – standard deviation, DNR – do not resuscitate advanced directive, <sup>a</sup>-MDS Item A1500, <sup>b</sup>-Study period is during vaccination season so may not represent the final seasonal vaccination rate, <sup>c</sup>-4 point score, 4 is severe impairment, e.g. comatose, <sup>d</sup>-any order for no resuscitation, chest compressions, <sup>e</sup>- MDS item J1400, <sup>f</sup>- for each resident how many times they had been tested in prior window |  |  |  |  |

**Table 2.** Cumulative incidence, event counts and effectiveness of booster dose versus nonreceipt

|  |  |  |  |  |  |
| --- | --- | --- | --- | --- | --- |
| **System 1: Private nursing home provider (n=200 facilities)** |  |  |  |  |  |
|  |  | **unboosted**<br>**(n=8,538)** | **boosted**<br>**(n=5,672)** | **Unadjusted VE** | **Adjusted VE<sup>a</sup>** |
| Any infection | Total days follow-up | 156,429 | 225,566 | 44.7% | 50.4% |
|  | Events (risk<sup>b</sup>) | 117 (0.75) | 77 (0.34) | (25.5%, 59.7%) | (29.4%, 64.7%) |
| Hospitalization | Total days follow-up | 159,177 | 227,243 | 32.7% | 47.7% |
|  | Events (risk<sup>b</sup>) | 5 (0.03) | 7 (0.03) | (-343.7%, 81.0%) | (-377.7%, 88.9%) |
| Death | Total days follow-up | 159,177 | 227,243 | 96.4% | 97.2% |
|  | Events (risk<sup>b</sup>) | 15 (0.09) | 1 (<0.01) | (84.8%, 100.0%) | (88.1%, 100.0%) |
| Combined | Total days follow-up | 159,177 | 227,243 | 78.6% | 82.0% |
|  | Events (risk<sup>b</sup>) | 20 (0.13) | 8 (0.04) | (52.1%, 93.0%) | (55.5%, 94.0%) |
| **System 2: VA Community-Living Centers (n=127 facilities)** |  |  |  |  |  |
|  |  | **unboosted**<br>**(n=4,100)** | **boosted**<br>**(n=2,291)** | **Unadjusted VE** | **Adjusted VE<sup>a</sup>** |
| Any infection | Total days follow-up | 81,538 | 92,665 | 69.3% | 58.2% |
|  | Events (risk<sup>b</sup>) | 73 (0.90) | 22 (0.24) | (52.1%, 82.5%) | (32.3%, 77.8%) |
| Hospitalization | Total days follow-up | 82,423 | 92,989 | 63.1% | 36.6% |
|  | Events (risk<sup>b</sup>) | 25 (0.30) | 9 (0.10) | (25.1%, 84.8%) | (-35.4%, 77.3%) |
| Death | Total days follow-up | 83,129 | 93,257 | 58.5%<sup>c</sup> | 39.5%<sup>c</sup> |
|  | Events (risk<sup>b</sup>) | 3 (0.04) | 2 (0.02) |  |  |
| Combined | Total days follow-up | 82,423 | 92,989 | 65.9% | 45.8% |
|  | Events (risk<sup>b</sup>) | 28 (0.34) | 10 (0.11) | (32.8%, 85.8%) | (-15.5%, 79.1%) |

