## supplementary appendix for "Effectiveness of a SARS-CoV-2 mRNA vaccine booster dose for prevention of infection, hospitalization or death in two nation-wide nursing home systems"

**Note:**

**System 1**: Residents from a large private provider of long-term care including 200 facilities concentrated in the northeast.

**System 2**: Veteran residents from 127 community-living centers administered by the Veterans Healthcare Administration

### **Figure S1**. Timing of third booster dose vaccinations

**System 1**

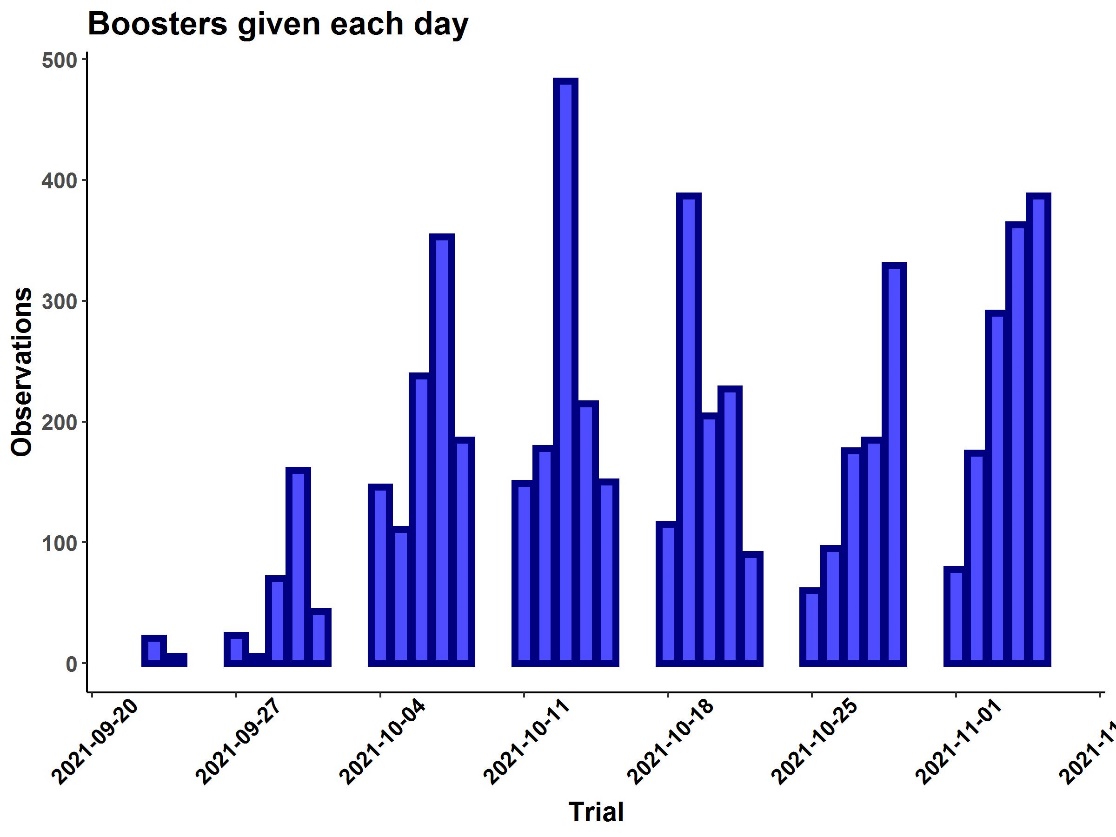

**System 2**

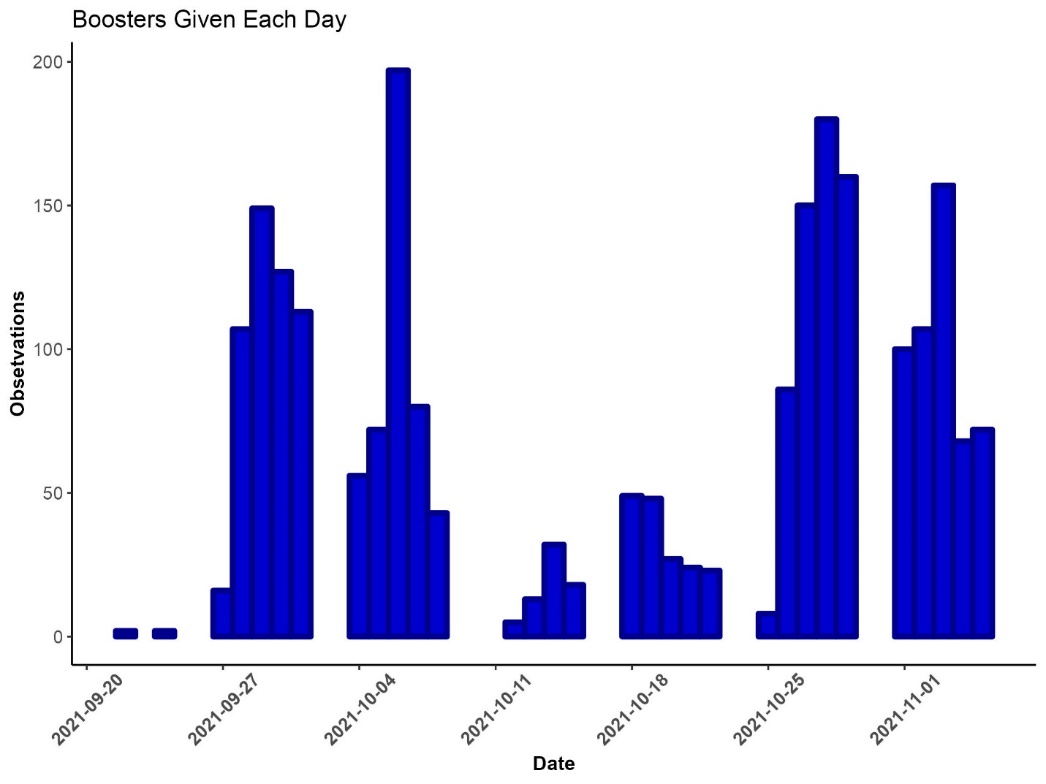

**Description.** Timing of booster vaccinations by calendar date.

### **Figure S3.** Exclusions over time

**System 1**

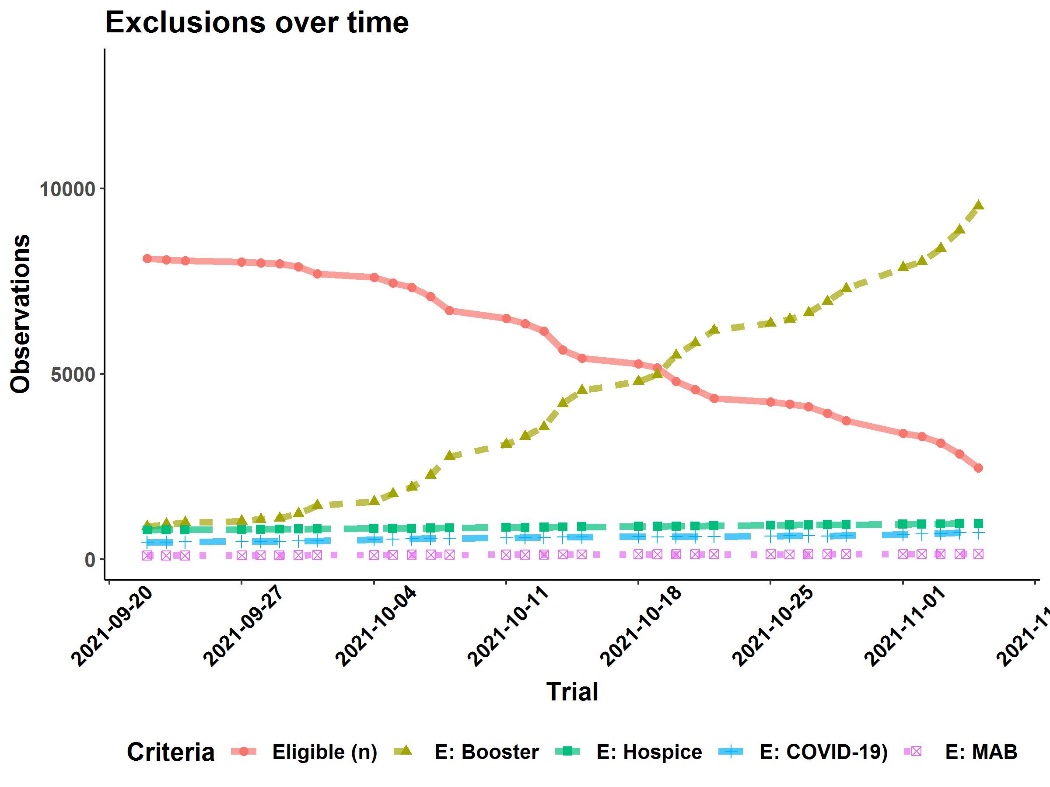

**System 2**
 
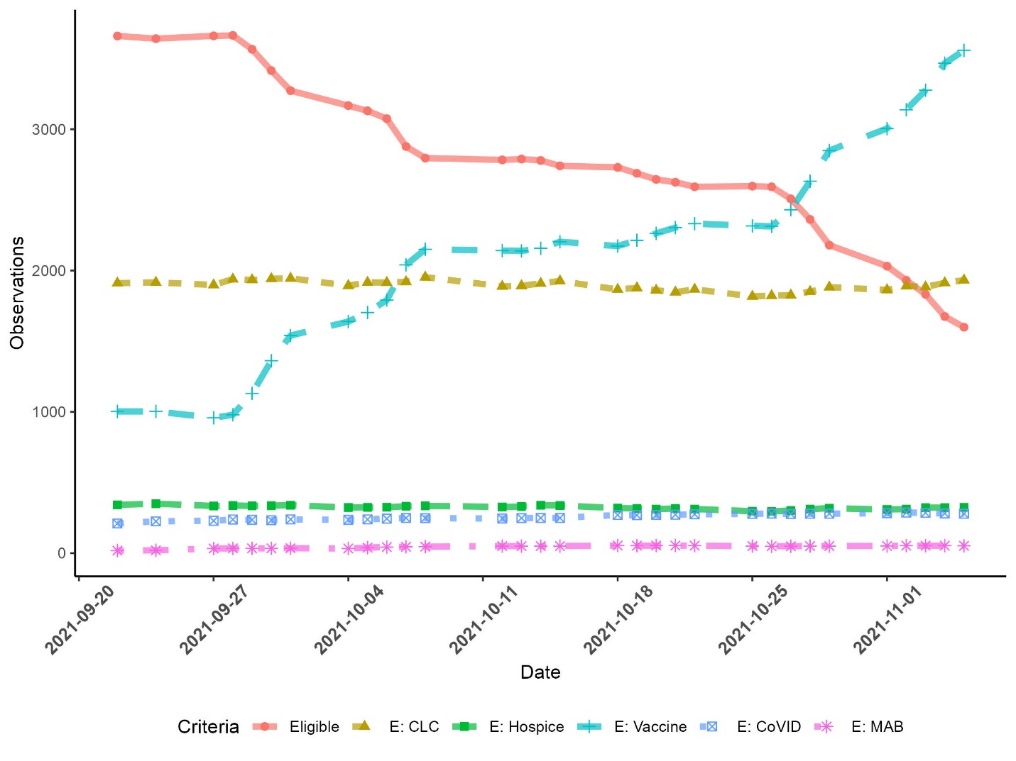

**Description**. Eligibility and reason for exclusion plotted over time. As more residents receive a third booster dose, this dominates reasons for non-eligibility.

### **Figure S4.** Propensity score overlap

**System 1**

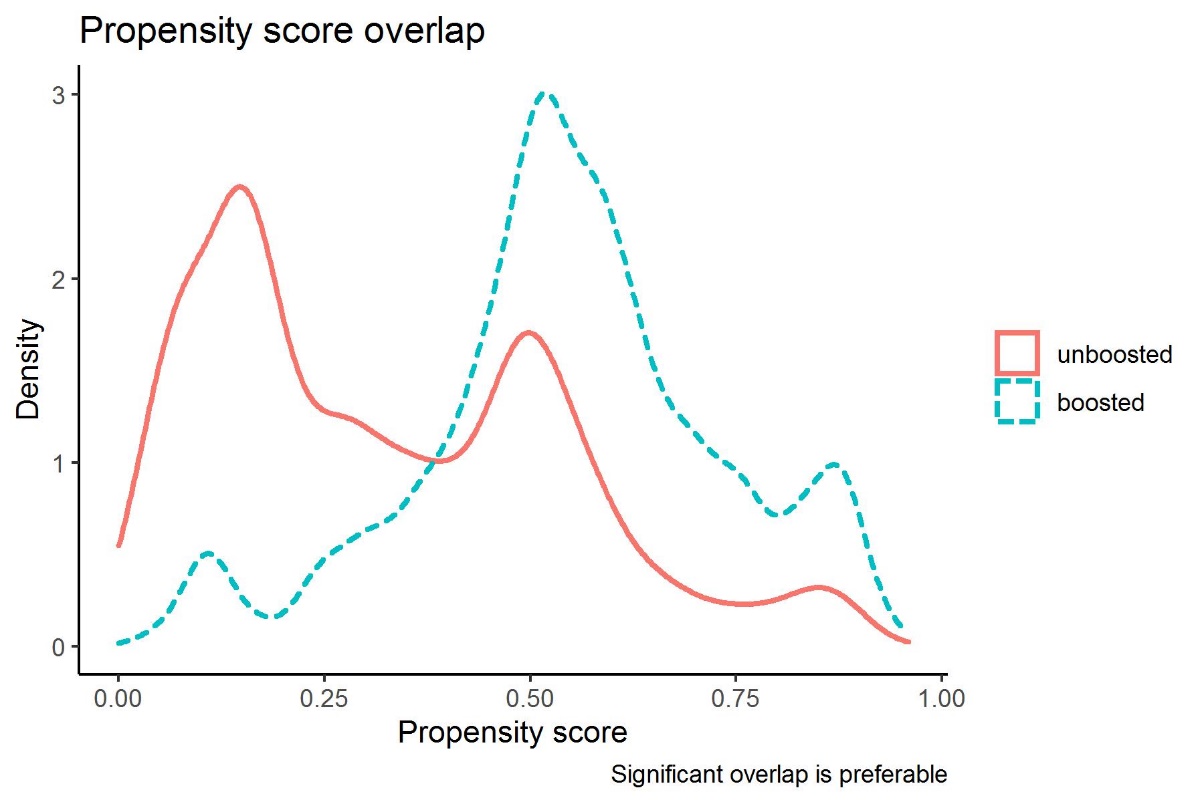

**System 2**

 
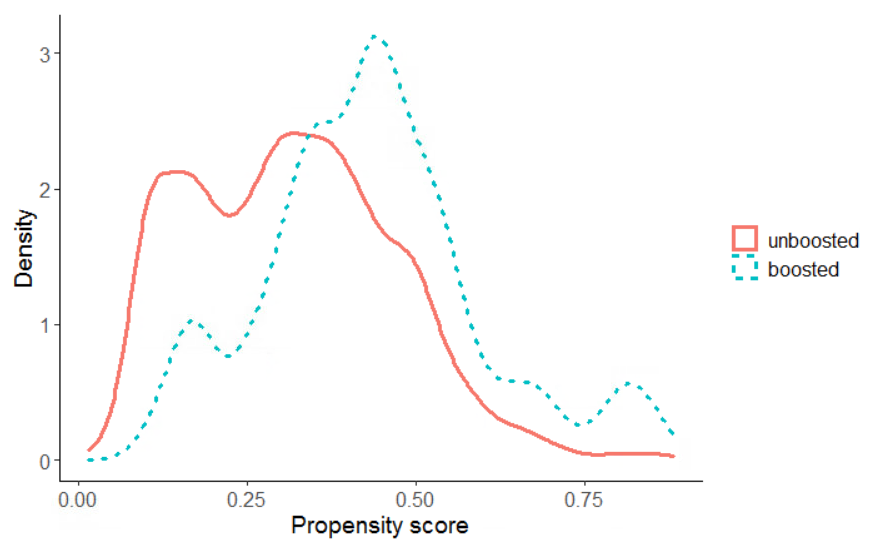

**Description.** Overlap of estimated propensity scores in boosted and non-boosted residents.

### **Figure S5**. Covariate balance pre- and post-probability weights

**System 1**

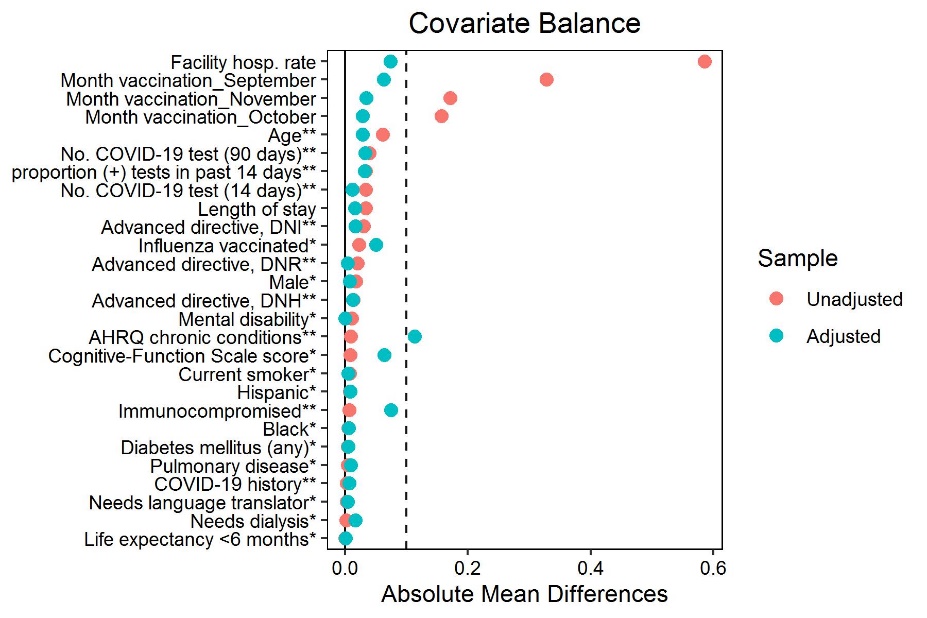

**System 2**

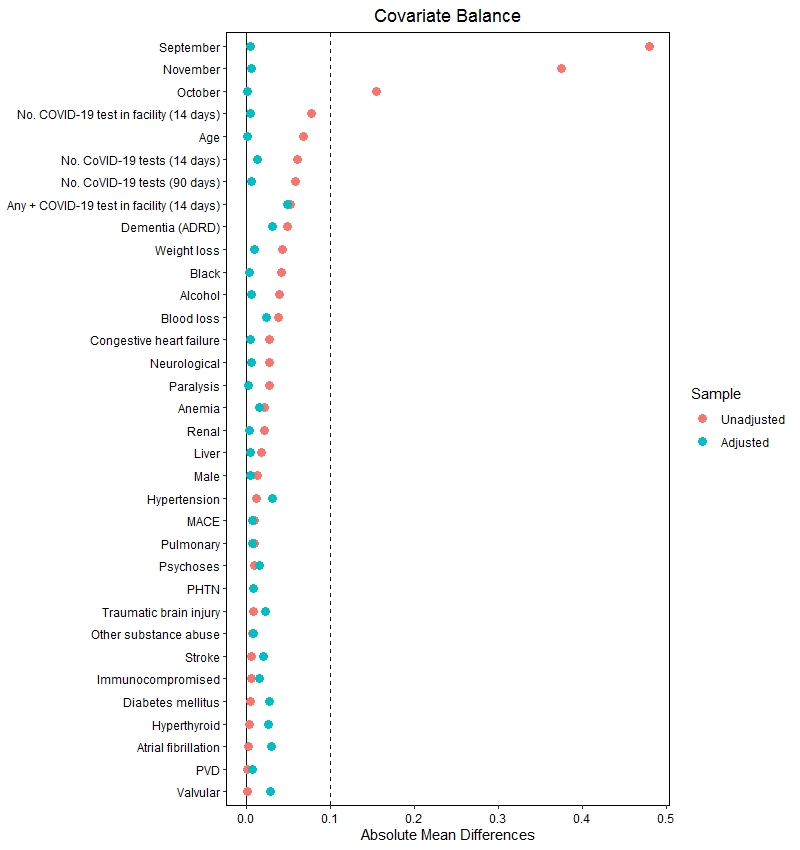

**Description**. The plot depicts the standardized mean differences (SMD) in boosted and non-boosted residents. The x-intercept line is at 0.1 SMD is given to reference a reasonable maximally acceptable difference between groups.

### **Table S1.** Definition of immunocompromised status

| **System 1 Immunocompromised definition** | |
| --- | --- |
| One of any of the following in the 6 months prior to the index date was used to classify individuals as immunocompromised | |
| **Medications** | **Diagnosis codes** |
| 5-fluorouracil, Abatacept, Adalimumab, Altretamine, Anakinra, Azathioprine  Basiliximab, Belatacept, Bendamustine, Budesonide, Busulfan  Carmustine, Certolizumab, Chlorambucil  Cisplatin, Cyclophosphamide, Cyclosporine, Calcineurin, Cytarabine  Diaziquone, Etanercept, Everolimus  Golimumab, Hydrocortisone, Hydroxychloroquine, Ixekizumab  Ifosfamide, Infliximab, Lomustine  Mechlorethamine, Melphalan, Mercaptopurine, Methotrexate, Mycophenolate  Natalizumab, Oxaliplatin,  Procarbazine, Rituximab  Secukinumab, Sirolimus, streptozocin  Tacrolimus, temozolomide  Thioguanine, thiotepa, Tocilizumab, Tofacitinib, Ustekinumab  Vedolizumab | **Immunocompromised status codes:** Z51.11, Z51.12, Z51.0, Z99.2, Z49.3, M35.9, L93.0  **Any code or branching code of the following:** Z94, Z49, B20, D61, D70, D71, D72, D73, D86, D89, C81, C82, C83, C84, C85, C86, C87, C88, C89, C90, C91, C92, C93, C94, C95, C96, D37, D38, D39, D40, D41, D42, D43, D44, D45, D46, D47, D48, M04, K50, K51, K52, M05, M06, M32, M34 |
| **System 2 Immunocompromised status** | |
| Abatacept, Abemaciclib, Acalabrutinib, Adalimumab , Ado-trastuzumab , Afatinib , Aldesleukin, Alectinib , Alefacept, Alemtuzumab , Alpelisib, Altretamine , Amifostine, Anakinra , Anti-thymocyte globulin, Apremilast , Arsenic, Asparaginase , Atezolizumab, Atezolizumab , Auranofin, Avapritinib , Avelumab, Aurothioglucose , Axicabtagene, Axitinib , Azacitidine, Azathioprine , Baricitinib, Basiliximab , Belatacept, Belimumab , Belinostat, Belumosudil , Bendamustine, Bevacizuma, Bexarotene, Binimetinib , Bleomycin, Blinatumomab , Bortezomib, Bosutinib , Brentuximab, Brigatinib , Brodalumab, Busulfan, Cabazitaxel, Cabozantinib, Calaspargase, Canakinumab, Capecitabine, Carboplatin, Carfilzomib, Carmustine, Cemiplimab, Ceritinib, Certolizumab, Cetuximab, Chlorambucil, Cisplatin, Cladribine, Clofarabine, Cobimetinib, Copanlisib, Crizotinib, Cyclophosphamide, Cyclosporine, Cytarabine, Dabrafenib, Dacarbazine, Daclizumab, Dacomitinib, Dactinomycin, Daratumumab, Dasatinib, Daunorubicin, Decitabine, Denileukin, Denosumab, Dimethyl fumarate, Dinutuximab, Diroximel fumarate, Docetaxel, Doxorubicin, Durvalumab, Eculizumab, Efalizumab, Elotuzumab, Emapalumab, Enasidenib, Encorafenib, Entrectinib, Enzalutamide, Epirubicin, Erdafitinib, Eribulin, Erlotinib, Estramustine, Etanercept, Etoposide, Everolimus, Fedratinib, Fingolimod, Floxuridine, Fludarabine, Fluorouracil, Flutamide, Fostamatinib, Fremanzumab, Gefitinib, Gemcitabine, Gemtuzumab, Gilteritinib, Glasdegib, Glatiramer, Glucarpidase, Golimumab, Guselkumab, Hydroxyurea, Ibritumomab, Ibrutinib, Idarubicin, Idelalisib, Ifosfamide, Imatinib, Infliximab, Inotuzumab, Inotuzumab, Interferon beta-1a, Interferon beta-1b, Ipilimumab, Irinotecan, Ivosidenib, Ixabepilone, Ixazomib, Ixekizumab, Lanadelumab, Lapatinib, Larotrectinib , Leflunomide, Lenalidomide , Lenvatinib, Lomustine, Lorlatinib, Lurbinectibdin, Mechlorethamine, Melphalan, Mercaptopurine, Methotrexate, Mitomycin , Mitotane, Mitoxantrone, Mogamulizumab, Mmuromonab, Mycophenolic acid, Natalizumab, Mycophenolate mofetil, Necitumumab, Nelarabine, Nilotinib, Nilutamide, Niraparib , Nivolumab, Obinutuzumab, Ocrelizumab, Ofatumumab, Olaparib, Olaratumab, Omacetaxine, Omalizumab , Osimertinib, Oxaliplatin, Paclitaxel, Palbociclib, Palivizumab, Panitumumab , Panobinostat, Pazopanib, Pegademase, Pegaspargase, Peginterferon/ribavirin, Peginterferon, Pembrolizumab, Pemetrexed, Pentostatin, Pertuzumab, Pexidartinib, Plicamycin, Pipobroman, Polatuzumab, Pomalidomide, Ponatinib, Pralatrexate, Procarbazine, Ramucirumab, Rasburicase, Ravulizumab, Regorafenib, Ribociclib, Rilonacept, Risankizumab, Rituximab, Romosozumab, Romidepsin, Rucaparib, Ruxolitinib, Sarilumab , Secukinumab, Selinex, Siltuximab, Siponimod, Sipuleucel, Sirolimus, vinblastine, Sonidegib, Sorafenib, Streptozocin, Sunitinib, Tacrolimus, Talazoparib, Talimogene, Tazemetostat , Temozolomide, Temsirolimus, Teniposide, Teriflunomide, Thalidomide, Thioguanine, Thiotepa, Tildrakizumab, Tildrakizumab, Tocilizumab, Tofacitinib, Topotecan, Tositumomab, Trabectedin, Trametinib , Trastuzumab, Trifluridine, Upadacitinib, Uracil mustard, Ustekinumab, Valrubicin, Vandetanib, Vedolizumab, Vemurafenib, Venetoclax, Vinblastine, Vincristine , Vinorelbine, Vismodegib , Voclosporin, Zanubrutinib | Any code or branching code of the following:    Z51.11, Z51.12, Z51.0, Z94, Z99.2, Z49, B20, D91, D70, D71, D72, D73, D75.81, D86, D89, C81, C82, C83, C84, C85, C86, C87, C88, C89, C90, C91, C92, C93, C94, C95, C96, D37, D38, D39, D40, D41, D42, D43, D44, D45, D46, D47, D48, M04, M05, M06, K50, K51, K52, M35.9, M32, L93.0, M34 |
| **Description.** The list of medications and diagnosis codes which were used to identify immunocompromised residents. System 2 includes a more comprehensive list developed and validated internally by Pharmacy Benefits Management for operational or research use in a variety of applications. It includes many medications that may be less relevant to a nursing home population, but is used for internal consistency in measurement of Veteran health outcomes across studies. | |

### **Table S2**. System 1 trial eligibility by date

| **Trial Date** | **Long-stay resident^a^** | **No discharge plan** | **Primary mRNA vaccination completed >=180 days prior** | **No history of SARS-CoV-2 in past 90 days** | **No history of monoclonal antibody use in past 90 days** | **Had not already received booster dose** | **Not currently receiving hospice care** |
| --- | --- | --- | --- | --- | --- | --- | --- |
| 9/22/2021 | 14562 | 12464 | 9346 | 9096 | 9062 | 8593 | 8117 |
| 9/23/2021 | 14557 | 12455 | 9345 | 9093 | 9059 | 8553 | 8080 |
| 9/24/2021 | 14544 | 12436 | 9342 | 9086 | 9052 | 8523 | 8054 |
| 9/27/2021 | 14442 | 12360 | 9313 | 9052 | 9018 | 8490 | 8022 |
| 9/28/2021 | 14408 | 12332 | 9299 | 9039 | 9003 | 8452 | 7989 |
| 9/29/2021 | 14361 | 12292 | 9281 | 9022 | 8986 | 8432 | 7971 |
| 9/30/2021 | 14318 | 12272 | 9266 | 9002 | 8966 | 8340 | 7886 |
| 10/1/2021 | 14259 | 12241 | 9238 | 8973 | 8932 | 8139 | 7699 |
| 10/4/2021 | 14123 | 12128 | 9188 | 8910 | 8871 | 8044 | 7607 |
| 10/5/2021 | 14078 | 12085 | 9177 | 8893 | 8855 | 7875 | 7448 |
| 10/6/2021 | 14047 | 12063 | 9172 | 8884 | 8846 | 7753 | 7334 |
| 10/7/2021 | 14011 | 12037 | 9159 | 8866 | 8828 | 7490 | 7081 |
| 10/8/2021 | 13977 | 12013 | 9145 | 8851 | 8811 | 7107 | 6712 |
| 10/11/2021 | 13859 | 11913 | 9105 | 8806 | 8766 | 6883 | 6501 |
| 10/12/2021 | 13830 | 11891 | 9101 | 8801 | 8759 | 6727 | 6348 |
| 10/13/2021 | 13787 | 11855 | 9076 | 8777 | 8736 | 6511 | 6150 |
| 10/14/2021 | 13745 | 11820 | 9051 | 8747 | 8704 | 5986 | 5646 |
| 10/15/2021 | 13715 | 11799 | 9031 | 8728 | 8686 | 5744 | 5420 |
| 10/18/2021 | 13618 | 11716 | 9026 | 8722 | 8679 | 5587 | 5264 |
| 10/19/2021 | 13572 | 11679 | 9038 | 8740 | 8694 | 5486 | 5163 |
| 10/20/2021 | 13533 | 11650 | 9057 | 8755 | 8712 | 5094 | 4793 |
| 10/21/2021 | 13492 | 11617 | 9025 | 8725 | 8682 | 4863 | 4574 |
| 10/22/2021 | 13456 | 11589 | 9004 | 8700 | 8657 | 4604 | 4336 |
| 10/25/2021 | 13364 | 11517 | 8984 | 8691 | 8649 | 4502 | 4242 |
| 10/26/2021 | 13337 | 11491 | 8976 | 8677 | 8636 | 4433 | 4178 |
| 10/27/2021 | 13307 | 11470 | 9005 | 8708 | 8667 | 4366 | 4112 |
| 10/28/2021 | 13258 | 11440 | 8982 | 8691 | 8646 | 4167 | 3930 |
| 10/29/2021 | 13219 | 11413 | 8962 | 8667 | 8621 | 3953 | 3733 |
| 11/1/2021 | 13114 | 11318 | 8909 | 8602 | 8555 | 3591 | 3390 |
| 11/2/2021 | 13075 | 11286 | 8897 | 8588 | 8541 | 3507 | 3313 |
| 11/3/2021 | 13057 | 11269 | 8891 | 8580 | 8533 | 3322 | 3137 |
| 11/4/2021 | 13030 | 11247 | 8873 | 8553 | 8506 | 3011 | 2833 |
| 11/5/2021 | 12986 | 11201 | 8839 | 8524 | 8477 | 2624 | 2459 |
| **Description.** On each day, eligibility is assessed according to outlined criteria. ^a^-Present in the home on that day, and had been resident for 90+ days. | | | | | | | |

### **Table S3**. System 2 trial eligibility by date

| **Trial Date** | **Long-stay resident** | **90 days in a CLC** | **Primary mRNA Vaccination Completed >=180 days prior & not yet boosted** | **No History of SARS-CoV-2 in past 90 days** | **No history of monoclonal antibody use in past 90 days** | **Not currently receiving hospice care** |
| --- | --- | --- | --- | --- | --- | --- |
| 2021-09-22 | 6389 | 4478 | 3910 | 3793 | 3791 | 3666 |
| 2021-09-24 | 6381 | 4465 | 3900 | 3774 | 3772 | 3647 |
| 2021-09-27 | 6365 | 4467 | 3922 | 3792 | 3783 | 3666 |
| 2021-09-28 | 6413 | 4474 | 3923 | 3790 | 3781 | 3670 |
| 2021-09-29 | 6409 | 4475 | 3820 | 3690 | 3681 | 3572 |
| 2021-09-30 | 6415 | 4473 | 3650 | 3535 | 3526 | 3421 |
| 2021-10-01 | 6403 | 4458 | 3499 | 3386 | 3377 | 3279 |
| 2021-10-04 | 6337 | 4444 | 3377 | 3274 | 3265 | 3173 |
| 2021-10-05 | 6366 | 4450 | 3337 | 3234 | 3225 | 3135 |
| 2021-10-06 | 6377 | 4462 | 3278 | 3175 | 3166 | 3080 |
| 2021-10-07 | 6386 | 4466 | 3068 | 2971 | 2962 | 2882 |
| 2021-10-08 | 6406 | 4453 | 2979 | 2887 | 2878 | 2800 |
| 2021-10-12 | 6361 | 4473 | 2972 | 2877 | 2868 | 2787 |
| 2021-10-13 | 6368 | 4476 | 2979 | 2881 | 2872 | 2794 |
| 2021-10-14 | 6391 | 4482 | 2969 | 2869 | 2860 | 2784 |
| 2021-10-15 | 6398 | 4472 | 2926 | 2826 | 2817 | 2746 |
| 2021-10-18 | 6341 | 4475 | 2934 | 2816 | 2806 | 2735 |
| 2021-10-19 | 6354 | 4477 | 2892 | 2774 | 2764 | 2693 |
| 2021-10-20 | 6345 | 4485 | 2849 | 2730 | 2720 | 2649 |
| 2021-10-21 | 6340 | 4493 | 2828 | 2712 | 2702 | 2629 |
| 2021-10-22 | 6349 | 4482 | 2790 | 2679 | 2669 | 2597 |
| 2021-10-25 | 6308 | 4491 | 2794 | 2681 | 2671 | 2601 |
| 2021-10-26 | 6312 | 4490 | 2792 | 2678 | 2668 | 2597 |
| 2021-10-27 | 6319 | 4493 | 2704 | 2592 | 2582 | 2511 |
| 2021-10-28 | 6343 | 4492 | 2542 | 2442 | 2432 | 2366 |
| 2021-10-29 | 6364 | 4482 | 2354 | 2256 | 2246 | 2183 |
| 2021-11-01 | 6345 | 4482 | 2195 | 2096 | 2086 | 2033 |
| 2021-11-02 | 6370 | 4480 | 2091 | 1993 | 1983 | 1933 |
| 2021-11-03 | 6369 | 4485 | 1984 | 1890 | 1880 | 1834 |
| 2021-11-04 | 6386 | 4473 | 1822 | 1730 | 1720 | 1677 |
| 2021-11-05 | 6388 | 4456 | 1747 | 1655 | 1645 | 1601 |

### **Table S4.** Baseline characteristics by booster status for VA nursing facilities (System 2)

| **Variable** | **Total**  (n=6,391) | **Unboosted** (n=4,100**)** | **Boosted**  (n=2,291) | **aSMD** |
| --- | --- | --- | --- | --- |
| Resident SARS-CoV-2 tests (past 14 days) | 1.86 (1.67) | 1.89 (1.67) | 1.79 (1.66) | 0.06 |
| Resident SARS-CoV-2 tests (past 90 days) | 11.39 (8.51) | 11.57 (8.48) | 11.08 (8.55) | 0.06 |
| Facility, # SARS-CoV-2 tests (past 14 days) | 146.81 (141.74) | 142.85 (137.86) | 153.90 (148.20) | 0.08 |
| Facility, any SARS-CoV-2 (+) in prior 14 days | 0.74 (1.96) | 0.70 (1.78) | 0.81 (2.25) | 0.05 |
| Length of stay in days, median (Q1, Q3) | 664  (338,1267) | 651  (341,1245) | 682  (380,1298) | 0.04 |
| Age, median (Q1, Q3) | 72 (68,78) | 72 (67,78) | 72 (68,78) | 0.07 |
| Male, no. (%) | 6,153 (96.28%) | 3,944 (96.20%) | 2,209 (96.42%) | 0.01 |
| White, no. (%) | 4,256 (66.59%) | 2,700 (65.85%) | 1,556 (67.92%) | 0.04 |
| Black, no. (%) | 1,581 (24.74%) | 1,040 (25.37%) | 541 (23.61%) | 0.04 |
| Race - Other , no. (%) | 554 (8.67%) | 360 (8.78%) | 194 (8.47%) | 0.01 |
| Immunocompromised, no. (%) | 1,865 (29.18%) | 1,193 (29.10%) | 672 (29.33%) | 0.01 |
| Current smoker, no. (%) | 707 (11.06%) | 486 (11.85%) | 221 (9.65%) | 0.07 |
| Pulmonary HTN | 209 (3.27%) | 136 (3.32%) | 73 (3.19%) | 0.01 |
| PVD | 1,635 (25.58%) | 1,049 (25.59%) | 586 (25.58%) | <.01 |
| Paralysis | 1,500 (23.47%) | 979 (23.88%) | 521 (22.74%) | 0.03 |
| Diabetes | 3,149 (49.27%) | 2,017 (49.20%) | 1,132 (49.41%) | <.01 |
| HTN | 4,325 (67.67%) | 2,782 (67.85%) | 1,543 (67.35%) | .02 |
| CHF | 1,464 (22.91%) | 956 (23.32%) | 508 (22.17%) | 0.03 |
| Pulmonary disease | 1,998 (31.26%) | 1,288 (31.41%) | 710 (30.99%) | 0.01 |
| Valvular disease | 491 (7.68%) | 315 (7.68%) | 176 (7.68%) | <.01 |
| Alcohol abuse disorder | 520 (8.14%) | 349 (8.51%) | 171 (7.46%) | 0.04 |
| Other substance abuse | 400 (6.26%) | 259 (6.32%) | 141 (6.15%) | 0.01 |
| Anemia | 2,397 (37.51%) | 1,553 (37.88%) | 844 (36.84%) | 0.02 |
| Malignancy | 739 (11.56%) | 469 (11.44%) | 270 (11.79%) | 0.01 |
| Psychoses diagnosis | 2,397 (37.51%) | 1,544 (37.66%) | 853 (37.23%) | 0.01 |
| Renal disease | 951 (14.88%) | 621 (15.15%) | 330 (14.40%) | 0.02 |
| Dementia (ADRD) | 4,275 (66.89%) | 2,709 (66.07%) | 1,566 (68.35%) | 0.05 |
| Atrial Fibrillation | 600 (9.39%) | 384 (9.37%) | 216 (9.43%) | <.01 |
| Traumatic Brain Injury | 461 (7.21%) | 293 (7.15%) | 168 (7.33%) | .01 |
| MACE | 2,468 (38.62%) | 1,590 (38.78%) | 878 (38.32%) | .01 |
| Stroke | 1,847 (28.90%) | 1,181 (28.80%) | 666 (29.07%) | <.01 |
| Hypothyroid | 892 (13.96%) | 574 (14.00%) | 318 (13.88%) | <.01 |
| Liver | 590 (9.23%) | 386 (9.41%) | 204 (8.90%) | 0.02 |
| Lymphoma | 73 (1.14%) | 44 (1.07%) | 29 (1.27%) | 0.02 |
| Metastasizing cancer | 134 (2.10%) | 90 (2.20%) | 44 (1.92%) | 0.02 |
| Rheumatic disease | 189 (2.96%) | 121 (2.95%) | 68 (2.97%) | <.01 |
| Coagulopathy, other | 413 (6.46%) | 265 (6.46%) | 148 (6.46%) | <.01 |
| Obesity | 988 (15.46%) | 641 (15.63%) | 347 (15.15%) | 0.01 |
| Recent weight loss | 905 (14.16%) | 602 (14.68%) | 303 (13.23%) | 0.04 |
| Blood loss | 184 (2.88%) | 127 (3.10%) | 57 (2.49%) | 0.04 |
| **Description**. aSMD – absolute standardized mean difference, Q1/Q3 – 25/75^th^ quartiles, SD – standard deviation. HTN – hypertension, CHF – congestive heart failure, PVD – peripheral vascular disease, ADRD – Alzheimer’s and related dementia. MACE – Major adverse cardiovascular event (i.e. MI, Stroke). Diagnoses determined from Elixhauser clinical condition categories. | | | | |
